# Systematic recovery of building plumbing-associated microbial communities after extended periods of altered water demand during the COVID-19 pandemic

**DOI:** 10.1101/2022.01.17.22269440

**Authors:** Solize Vosloo, Linxuan Huo, Umang Chauhan, Irmarie Cotto, Benjamin Gincley, Katherine J Vilardi, Byungman Yoon, Kelsey J Pieper, Aron Stubbins, Ameet Pinto

## Abstract

Building closures related to the coronavirus disease (COVID-19) pandemic resulted in increased water stagnation in commercial building plumbing systems that heightened concerns related to the microbiological safety of drinking water post re-opening. The exact impact of extended periods of reduced water demand on water quality is currently unknown due to the unprecedented nature of widespread building closures. We analyzed 420 tap water samples over a period of six months, starting the month of phased reopening (i.e., June 2020), from sites at three commercial buildings that were subjected to reduced capacity due to COVID-19 social distancing policies and four occupied residential households. Direct and derived flow cytometric measures along with water chemistry characterization were used to evaluate changes in plumbing-associated microbial communities with extended periods of altered water demand. Our results indicate that prolonged building closures impacted microbial communities in commercial buildings as indicated by increases in microbial cell counts, encompassing greater proportion cells with high nucleic acids. While flushing reduced cell counts and increased disinfection residuals, the microbial community composition in commercial buildings were still distinct from those at residential households. Nonetheless, increased water demand post-reopening enhanced systematic recovery over a period of months, as microbial community fingerprints in commercial buildings converged with those in residential households. Overall, our findings suggest that sustained and gradual increases in water demand may play a more important role in the recovery of building plumbing-associated microbial communities as compared to short-term flushing, after extended periods of altered water demand that result in reduced flow volumes.

## Introduction

Social distancing policies enacted in March 2020 during the initial wave of the coronavirus (COVID-19) pandemic to combat disease spread^1,2^ caused widespread building closures in the non-residential sector (i.e., commercial, industrial, etc.). These measures limited building occupancy and adjusted individual and societal behaviors (e.g., homebased remote learning and working practices, hygiene practices)^3^, which significantly altered water demand at the water distribution and building system levels^4–10^. The impact of policy interventions related to the COVID-19 pandemic on water demand have recently been addressed^4–10^. Several studies have reported significant reductions in water demand across non-residential sector buildings^5,8–10^. For example, Li et al. (2020) reported a decrease of 11.6% in water demand across non-residential sectors in California (United States), while Kalbusch et al. (2020) documented reductions in water demand, ranging between 30% and 53%, across non-residential sectors in Joinville, Brazil. Contrasting, reports in residential sectors (houses, apartments, and condominiums) indicate increases in water demand^6–9^, as well as anomalies in water demand peaks^7,10^.

Reduced water demand resulting in lower flow volumes and higher stagnation times in the building plumbing have been linked to water quality deterioration and several issues that are of public health concern, including microbial regrowth^11–14^, opportunistic plumbing pathogen (OPP) growth^12,15– 17^, metal (e.g., lead) leaching^18^, and disinfectant byproduct formation (trihalomethanes (THMs), haloacetic acids (HAAs))^19^. Unlike drinking water distribution systems, building plumbing systems experience significant loss in disinfectant residual, have large temperature gradients and, consist of unique site-specific features (e.g., heterogenous materials, high surface area to volume ratios, onsite storage tanks) that impact water quality and promote microbial and OPP growth during periods of low water demand^14,17,20^. In contrast to drinking water distribution systems that are subject to water monitoring plans, building plumbing systems rely on guidance documents, consisting of remediation actions, to ensure safe water quality at point of use (PoU)^21^. Remedial actions recently incorporated in guidance documents to minimize risks associated with water quality deterioration in buildings impacted by COVID-19 social distancing policies, include continuous or shock disinfection, routine flushing, and diagnostic testing of OPPs (*Legionella* spp.)^21,22^. The scope and details of these remedial actions varies widely and depend on site-specific factors, i.e., building size and plumbing configuration, as well as the risk(s) that needs to be mitigated. Flushing replenishes disinfectant residual concentrations as well as reduces microbial and OPP loads in building plumbing systems^11,14,16,17^. The effectiveness of flushing practices in restoring water quality to baseline conditions are typically assessed by performing intermittent testing of temperature and disinfectant residual concentrations^21^. Stabilization of these measures during flushing are indicative of baseline water quality conditions^13^; however, neither of these measures provide direct microbiological insight. Flow cytometry has been an established technology for high-resolution monitoring of microbial communities in water system^23^ as it provides direct quantitative measures of cell concentrations as well shifts in microbial community fingerprints^24–27^. For example, it has been used to monitor pipe flushing after overnight stagnation in buildings^11,13^ and during maintenance operations in the drinking water distribution systems^28^.

Managing water quality in buildings is crucial to avoid plumbing-associated health risks; however, consensus in guidance documents and expert opinions on design and operational topics that are critical for water quality management are lacking^21,22^. Data generated from field studies can drive the development of evidence-based best practices for water quality management, which can facilitate more informed and coherent decisions. This study examines the dynamics of plumbing-associated microbial communities in response to altered water demand during the COVID-19 pandemic in Boston, Massachusetts. Drinking water samples were collected monthly over a period of six months, starting the month of phased reopening, i.e., June 2020, from commercial buildings and residential households and characterized using flow cytometry as well as a comprehensive suite of water chemistry parameters. Our objectives were (1) to evaluate the impact of extended periods of altered water demand on microbial communities, (2) to assess the impact of flushing on drinking water microbial communities, and (3) to determine the impact of phased reopening on the drinking water microbial communities in residential and commercial building plumbing.

## Materials and Methods

### Description of study site and sample collection

Shortly after a state of emergency was declared in Massachusetts (MA) (March 10, 2020), college campuses across MA transitioned to remote learning. As a result, classrooms, residence halls, and assembly buildings (e.g., dinning services) were closed and non-essential research operations were halted on multiple campuses including Northeastern University (NEU) in Boston. On June 1^st^, NEU partially reopened for research activities at established laboratory capacity limits of 25% for the remainder of the year. For the Fall 2020 semester, residence halls and classrooms opened at reduced capacity and NEU operated with hybrid learning, which encompassed part in-person and part remote learning. In this study, three NEU buildings (hereafter, commercial buildings) and four residential households were monitored to evaluate the impact of varying water demand on building plumbing-associated microbial communities. These site types (commercial buildings and residential households) (i) ranged in size, age, and functionality, (ii) were situated within five miles of each other and (iii) were served with chloraminated water from the same distribution system. Cold water taps at each site type, i.e., commercial building (number of sites =3, number of taps per site = 2) and residential households (number of sites = 4, number of taps per site = 1), were selected for monthly sampling over six months. These taps included taps from residential kitchens (*n* = 2), commercial kitchenets (*n* = 3), residential bathrooms (*n* = 2), and research laboratories in commercial buildings (*n* = 3). Flushing profiles were conducted at all ten taps monthly on two consecutive days. Seven samples were obtained from each tap including the first draw sample (time point 1 (TP1, 0 min)) and six flushed samples that were collected at five min intervals over a period of 30 min (TP2, 5 min; TP3, 10 min; TP4, 15 min, TP5, 20 min, TP6, 25 min, and TP7, 30 min). Samples collected at TP1 and TP7 included a 2,000 mL sample in 2 L narrow-mouth polycarbonate bottles (Thermo Scientific™, Cat. No.: DS22050210) as well as a 500 mL sample in 500 mL wide-month HDPE bottles (Thermo Scientific™, Cat. No.: 02-896-2E). The latter sample were included for metal analysis, while aliquots (50 mL sample) of the 2,000 mL sample were used for measures of water chemistry. Post-first flush samples (i.e., TP2 – TP6) were collected in sterile 15 ml polyethylene centrifuge tubes (Falcon™, Cat No.: 352196). Intermittent temperature measurements were obtained throughout the duration of sampling at 10 sec intervals using an Elitech GSP-6 data logger and flow rates averaged at 4.10 ± 1.80 L.min^−1^, ranging between 2.00 and 8.19 L.min^−1^ (Table S1).

### Water chemistry analysis

Water quality parameters (i.e., temperature, pH, conductivity, and dissolved oxygen) were measured using the Orion Star™ A325 pH/Conductivity Portable Multiparameter Meter (Thermo Scientific™, Cat. No.: STARA3250). Total chlorine was measured using HACH Method 8167 with DPD Total Chlorine Reagent Powder Pillows (HACH, Cat. No.: 2105669). Nitrogen species, including ammonium, nitrate, and nitrite were measured using the Nitrogen-Ammonia Reagent Set (Method 10023, HACH, Cat. No.: 2604545), NitraVer X Nitrogen-Nitrate Reagent Set (Method 10020 HACH, Cat. No.: 2605345), and NitriVer 3 TNT Reagent Set (Method 10019, Cat. No.: 2608345), respectively. All HACH measurements were performed on the DR1900 Portable Spectrophotometer (HACH, Cat. No.: DR190001H). For metal analysis, samples were acidified with 2% nitric acid and digested from a minimum of 16 hours before analysis on the Inductively Coupled Plasma – Mass Spectrometry (ICP-MS) according to method 3125 B^29^. Blanks and spikes of known concentrations were measured every 10 samples for all analyses for quality control. Samples for dissolved organic carbon (DOC) and total dissolved nitrogen (TDN) were acidified to pH < 2 using HCl (p.a.) before analysis using a Shimadzu TOC-VCPH total organic carbon analyzer with a Total Nitrogen Module (TNM) attached^30^. Certified DOC and TDN quality references from Ultra Scientific (QCI-731 and QCI-745Agilent) were measured to confirm precision and accuracy. Measured DSR values were consistent with the consensus values with a standard deviation <5%. Routine minimum DOC detection limits using the above configuration are 0.034 ± 0.004 mg L^−1^ and standard errors are typically 1.7 ± 0.5 % of the DOC concentration^30^. For TDN, routine minimum detection limits are 0.041 ± 0.003 mg L^−1^ and standard errors are typically 0.8 ± 0.4 % of the TDN concentration^30^.

### Flow cytometry analysis

Samples were processed in triplicate to obtain flow cytometric (FCM) measurements of total cell concentrations (TCC) and intact cell concentrations (ICC) as described previously^31–33^. Briefly, samples (500 μl) treated with 10mM sodium thiosulfate (1% (v/v)) (Alfa Aesar™, Cat. No.: AA35645K2) were pre-heated at 37°C for 3 min and then stained with SYBR Green I (SG) (Invitrogen™, Cat. No.: S7585) (1:100 SG diluted in 10 mM Tris-HCl (pH 8.5, Bioworld, Cat. No: NC1213695) at 10 μL.ml^−1^ (TCC measurements) or SG combined with propidium iodide (PI) (Molecular Probes™, Cat. No.: P3566) (3 μM final concentration) at 12 μL.ml^−1^ (ICC measurements). After staining the samples were incubated in the dark at 37°C for 10 min using the Eppendorf ThermoMixer® 2C (Eppendorf, Cat. No.: 2231000680). Five negative controls were processed identically and in parallel with the samples, including (i) unstained UltraPure™ DNase/RNase-Free Distilled Water (Thermo Fisher Scientific, Cat. No.: 10977015), (ii) SG stained UltraPure™ DNase/RNase-Free Distilled Water, (iii) SGPI stained UltraPure™ DNase/RNase-Free Distilled Water, (iv) SG stained 0.22μm filtered drinking water sample, and SGPI stained 0.22μm filtered drinking water sample. FCM analysis was performed on 50 μL sample at a pre-set flow rate of 66 μl.min^−1^ using a BD Accuri® C6 flow cytometer (BD Accuri® cytometers, Belgium), which is equipped with a 50mW solid state laser emitting light at a fixed wavelength of 488 nm. Green and red fluorescent intensity were collected at FL1 = 533 ± 30 nm and FL3 > 670 nm, respectively, along with sideward (SSC) and forward (FCS) scatter light intensities. Flow Cytometry Standard (FCS) files were processed in R v4.1.2^34^ using the Phenoflow package v1.1^24^ and its dependencies (https://github.com/CMET-UGent/Phenoflow_package). Briefly, FCS files were imported into R using flowCore v2.2.0^35^ and then four parameters in signal height format (i.e., FL1-H, FL3-H, SSC-H, and FSC-H) were extracted and rescaled using hyperbolic arcsine transformations. A fixed polygonal gate was applied on the FL1-H and FL3-H graph to separate signal (i.e., cells) and background noise (i.e., instrument noise and (in)organic background)^36,37^ and to estimate measures of ICC and TCC as cells.μL^−1^. A threshold on FL1-H was selected to estimate high - and low nucleic acid cell concentrations in cells.μL^−1^ (HNA and LNA, respectively). The function flowBasis were used for advance fingerprinting, with nbin set to 128 and bw (band width) set to 0.01^24^. From this, biodiversity estimates, i.e., phenotypic diversity index (D2; arbitrary units, a.u.) and evenness (a.u.), were calculated as well as measures of beta diversity through Bray-Curtis dissimilarity using, in addition, the vegan package^38^. All biodiversity measures were performed after resampling to the sample with the lowest cell count. The raw FCS files have been deposited in FlowRepository and are publicly available under accession number FR-FCM-Z4SR.

### Statistical analysis

Statistical analysis was performed using packages in R v4.1.2^34^. Water quality parameters, i.e., water chemistry measures and flow cytometric measurements, were assessed for normality using the Shapiro-Wilks test provided in the stats package^34^ and then comparisons between groups were done by performing either independent t-test (t.test{stats}; parametric test for independent samples) or Mann-Whitney U test (wilcox.test{stats}; non-parametric test for independent samples). Non-multidimensional scaling (NMDS) was performed using metaMDS provided in the vegan package^38^ to visualize differences in beta diversity based on Bray-Curtis dissimilarity distances and permutational multivariate analysis of variance (PERMANOVA) was performed using adonis{vegan}. To identified localized factors driving change in the microbial community composition, water chemistry data were z-score standardized and considered for Bray-Curtis distance-based redundancy analysis (dbRDA) using dbrda{vegan}. To circumvent for bias associated with collinearity, colinear variables with significant Pearson correlation coefficient > 0.70 and < -0.70 were identified and then placed into clusters followed by the selection of one variable per cluster for dbRDA^39^. The excluded colinear variables were considered for discussion and placed in brackets, with “+” denoting positive correlations (Pearson correlation coefficient (R^2^) > 0.70) and “-” denoting negative correlations (R^2^ < -0.70). Significance of each response variable was confirmed with an ANOVA (anova{stats}) and only response variables with significance < 0.01 were kept in the model. The explanatory value of significant response variables was assessed with variation partitioning analysis using varpart{vegan}. All plots were generated using ggplot2^40^.

## Results and discussion

### Extended periods of altered water demand significantly impacted water quality metrics in commercial as compared to residential buildings

Monthly water demand for the commercial building and residential household sites from January 2019 to November 2020 are presented in Figure 1A. At both site types, water demand levels from January to February of 2020 corresponded to levels that were documented in 2019 (Table S2); however, starting March 2020, notable different water demand patterns were observed. Commercial building sites had low water demand levels between March and May 2020 (mean ± standard deviation (SD) = 12,928 ± 6,183 m^3^.month^−1^), which were 39% to 46% lower compared to 2019 (22,088 ± 9,287 m^3^.month^−1^) due to building closures and individuals transitioning to off-campus work and remote learning. These observations concur with recent studies that reported notable reductions in water demand across buildings in commercial, industrial, and institutional sectors, due to COVID-19 social distancing policies^5,8–10^. In contrast, the residential sites were associated with water demand increases between March and May 2020, which agrees with recent studies^6–9^. These ranged between 743 and 1,987 m^3^.month^−1^, with an average 6% increase in water demand compared to 2019 (1,245 ± 295 m^3^.month^−1^; Table S2) and were due to social distancing policies that prompted “stay at home” orders, which facilitated remoted learning and office practices, as well as added hygienic actions (e.g., increased hand washing)^1–3^.

**Figure 1.**
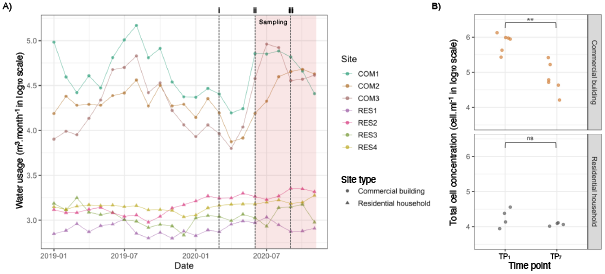
**A)** Water demand in cubic meter per month associated with three commercial buildings (COM, •) and four residential household (RES, ▴) sites from January 2019 – November 2020. The dashed lines indicate: i, closure of commercial building sites due to COVID-19 policy interventions; ii, reopening of commercial building sites for research activity; and iii, reopening of commercial building sites for the Fall 2021 academic year. The red shaded area indicates the sampling period of six months (June to November 2020). **B)** Total cell concentration measured in cells per ml (averaged over triplicate measurements) for first draw (TP1) and final flushed (TP7) samples that were collected in the commercial building (orange) and residential household (grey) sites the first month of sampling following phased reopening (i.e., June). Differences between TP1 and TP7 of the commercial building and residential household sites were assessed using the Mann-Whitney U test and significance are indicated with a significant bar, where 0.01 ≤ p < 0.05 (*), 0.001 ≤ p < 0.01 (**), p < 0.001 (***), and not significant (ns).

Low water demand at the commercial building sites impacted water quality due to elevated water stagnation^22^. Under normal building operations, stagnation-induced effects over multiple time scales (i.e., overnight, over weekends) have been linked to water quality deterioration that relate to microbial regrowth^11–14^, opportunistic plumbing pathogen (OPP) growth^12,15–17^, and metal (e.g., lead) leaching^18^. In this study, the premise plumbing systems of the commercial building sites had reduced flow volumes resulting in stagnation for approximately 14 weeks (March 13 – June 4, 2020) and presented evidence of impacted water quality (Table S3A). Total cell concentration (TCC) measures of the samples that were collected from the commercial building sites during the first month of phased reopening (i.e., June 2020) averaged at 2.95 ± 3.64 × 10^5^ cells.ml^−1^ (range = 1.19 × 10^4^ to 1.34 × 10^6^ cells.ml^−1^) for the entire flushing period and were significantly higher when compared to the residential household sites (mean ± SD = 1.11 ± 0.58 × 10^4^ cells.ml^−1^; Mann-Whitney U test, p < 0.05; Table S3A). The first draw sample (TP1) collected from both site types were associated with higher TCC measures than the final flushed samples (TP7; Figure 1B). Specifically, differences in TCC measures of the commercial building TP1 samples (8.07 ± 3.68 × 10^5^ cells×mL^−1^) were significant and approximately 8-fold higher when compared to the TP7 samples (1.00 ± 0.89 × 10^5^ cells×mL^−1^; p < 0.05; Table S3B), while smaller insignificant 2-fold differences in TCC measures were observed between the TP1 and TP7 samples of the residential household sites (2.07 ± 1.09 × 10^4^ and 1.18 ± 0.12 × 10^4^ cells×mL^−1^, respectively; p > 0.05; Table S3B). The microbial loads at the commercial building samples were associated with high temperatures, low total chlorine concentrations, and elevated levels of metals, particularly copper and manganese that were significantly different when compared to the residential household sites (all p < 0.05; Table S3A). High microbial loads, coinciding with low residual disinfectant concentrations and elevated temperatures were in consensus with previous studies that were conducted over shorter time scales (i.e., overnight, over weekends)^11,13–15,41^. These findings confirm the impact of reduced water demand that led to stagnation and impacted water quality^21,22,42^.

The samples collected from the commercial building sites had greater intact cell proportions, averaging at 29.81 ± 20.24% and were associated with less diverse (D2 = 2126 ± 350) and even (evenness = 0.23 ± 0.02) microbial communities, compared to the residential household samples (Figure 2A Panel A) (Mann-Whitney U test, all p < 0.05; Table S3A). Differences in intact cell proportions of the commercial building TP1 samples (52.98 ± 3.42%) were significant and approximately 2-fold higher than the TP7 samples (24.6 ± 14.22%) (p < 0.05; Table S3B). The TP1 samples were also associated with less diverse and even microbial communities, which were significant when compared to the TP7 samples (Figure 2A Panel B and C) (independent t-test, all p < 0.05; Table S3B). Similar but to a lesser extent insignificant differences in intact cell proportion, D2, and evenness were observed between the TP1 and TP7 samples of the residential household sites (Figure 2A Panel B and C) (Mann-Whitney U test or independent t-test, all p > 0.05; Table S3B). Further exploration of microbial community composition using structure-based Bray-Curtis dissimilarity distances showed clear clustering of samples based on site type (Figure 2B), which explained approximately 15% of the variation in the microbial community composition (PERMANOVA, F(1, 208) = 37.00, R2 = 0.151, p < 0.05). Localized water chemistry parameters driving change in the microbial community composition of the commercial building and residential household sites were identified using Bray-Curtis dbRDA (Figure 2C). Variability in microbial community composition was significantly explained by site type and six water chemistry parameters, including temperature, dissolved oxygen [-Pb^208^], total chlorine [-Cu^65^], ammonium, Mn^55^ [+Cu^65^, +Mg^24^], and Zn^66^ (permutation test for Bray-Curtis dbRDA, all *p* < 0.01; Table S4A and S4B). The dbRDA biplot presented in Figure 2C, shows clear separation of samples by site type that explained approximately 12% of the variation in microbial community composition based on variance partitioning analysis (adj. R^2^ = 0.119; Table S4C). Furthermore, total chlorine [-Cu^65^], ammonium, and dissolved oxygen [-Pb^208^] were identified as primary water chemistry parameters associated with change in microbial community composition between site types (variance partitioning analysis, adj. R^2^ = 0.115, 0.088, and 0. 029; Table S4C). Of these, total chlorine explained most of the variation ∼12% and was present at significantly lower concentrations in the commercial building samples (0.51 ± 0.68 mg.l^−1^) compared to the residential household samples (1.57 ± 0.6 mg.l^−1^; Mann-Whitney U test, p < 0.05; Table S3A). Loss of disinfectant residual during intermittent periods of stagnation have been shown to drive change in plumbing-associated microbial community compositions^14,20,41^. Further, the commercial building sites were associated with higher Mn^55^, Mg^24^, Zn^66^, Cu^65^ and Pb^208^ concentrations, which likely relates to metal leaching from the piping materials or from scale formation during stagnation and subsequent release^43^. All measured metals concentrations were consistently below the regulatory concentrations.

**Figure 2:**
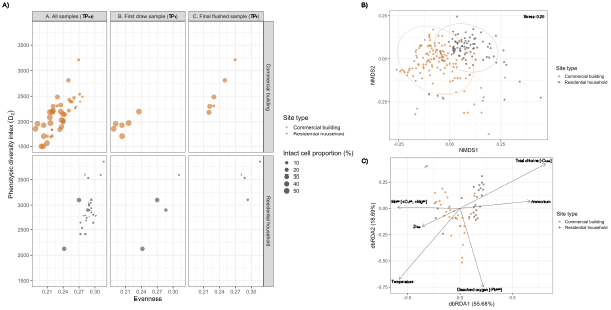
**A)** Bubble plot showing the proportion intact cells (depicted by size) with phenotypic diversity index (D2) and evenness of TP1-7 samples that were collected the first month of phased reopening from commercial building (orange) and residential household (grey) sites. Panels from the left to right represents measures related to (i) all samples (TP1-7, Panel A), (ii) first draw samples (TP1, Panel B), and (iii) final flushed samples (TP7, Panel C). **B)** NMDS plot illustrating differences in microbial community composition between TP1-7 samples of commercial buildings (orange) and residential households (grey) sites, using Bray-Curtis dissimilarity distances on flow cytometric data (i.e., FL-1, FL-3, FCS, and SSC). Ellipse are drawn at 95% confident limit. **C)** Bray-Curtis distance-based redundancy analysis (dbRDA) biplot illustrating the relationship between site type, water chemistry parameters and microbial community composition of TP1 and TP7 samples of the commercial building (orange) and residential household (grey) sites. Water quality parameters that significantly explained variation in community composition based on permutational ANOVA for Bray-Curtis dbRDA (p < 0.01) are shown as black arrows and the percent variation explained by each axis is shown in parenthesis. Colinear factors are shown in brackets, with “+” denoting positive correlations (Pearson correlation coefficient (R^2^) > 0.70) and “-” denoting negative correlations (R^2^ < -0.70).

### Flushing profiles demonstrated differential impact of stagnation in commercial and residential buildings varied with phased re-opening

The first draw samples (TP1) that were collected from both site types had higher TCC measures than the flushed samples (TP2-7; Figure 3A). Specifically, TCC measures of the commercial building TP1 samples were significantly higher (>3-fold) when compared to the TP2-7 samples (Mann-Whitney U test, all p < 0.05, Table S5). In contrast, differences in TCC measures between the TP1 and TP2-7 samples of the residential household sites were less than 2-fold and largely insignificant (Mann-Whitney U test and independent t-test, p > 0.05; Table S5). Overall, these findings suggest that flushing practices counter for stagnation-induced effects at both site types. Further, flushing practices at the residential household sites more rapidly mitigated stagnation-induced effects as compared to the commercial building sites. Specifically, TCC measures of the residential household sites stabilized at concentrations ranging between 3.82 × 10^3^ cells.ml^−1^ and 1.99 × 10^4^ cells.ml^−1^, after flushing approximately 19.88 ± 6.12 L water in 5 min (Figure 3A, Table S6). Similar trends were observed with temperature measures that stabilized after an initial decrease between TP1 and TP2 (Figure S1). Depending on the sampling month, these temperature reductions ranged between 1°C and 4°C (Table S6). These congruent patterns imply that fresh water from the distribution main reached the point of use (PoU) fixtures within the first five minutes of flushing, and that stagnant water were removed from the plumbing systems of the residential household sites^13^. In contrast, TCC measures of the commercial building sites presented little evidence of stabilization and were associated with large differences between TP1 (TCC measures averaged across months = 5.12 ± 2.22 × 10^5^ cells.ml^−1^) and TP2 (1.30 ± 1.49 × 10^5^ cells.ml^−1^), while successive time points had smaller differences and continually decreased in sequential order (Figure 3A, Table S6). These changes were linked to elevated temperature measures that presented little to no evidence of stabilization and ranged between 20.40°C and 24.50°C (Figure S1, Table S6). Overall, these findings imply that fresh water from the distribution main did not reach the PoU fixtures of the commercial building sites during 30 min of flushing and that stagnation-induced effects in the plumbing systems were persistent. This difference between commercial buildings and residential households is not surprising and is a direct result of the size of the plumbing systems at the sampling sites.

**Figure 3:**
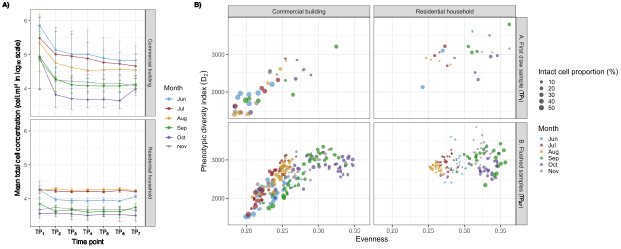
**A)** Monthly flush profiles based on total cell concentration (TCC) measurements that were averaged for individual time points (TP1, 0 min; TP2, 5 min; TP3, 10 min; TP4, 15 min; TP5, 20 min; TP6, 25 min; and TP7, 30 min) within each month (Jun, blue; Jul, red; Aug, yellow; Sep, green; Oct, purple; and Nov, grey) across all commercial building and residential household sites, respectively. Bars represent standard deviation, indicating the dispersion of individual TCC measures in relation to the mean. B) Bubble plot showing intact cell proportions (depicted by size) with phenotypic diversity index (D2) and evenness of monthly samples that were collected from the commercial building and residential household sites over six months. Panels from the top to bottom represents measures related to (i) first draw samples (TP1, Panel A), and (ii) all flushed samples (TP2-7, Panel B).

The microbial community composition of the TP_1_ samples were different from the TP_2-7_ samples and were associated with greater intact cell proportions as well as less diverse and even microbial communities (Figure 3B). These differences were significant and more noticeable at the commercial building sites; however, largely insignificant at the residential household sites (Mann-Whitney U test or independent t-test; Table S5). Structure-based Bray-Curtis dissimilarity distances indicated significant differences between TP_1_ and TP_2-7_ samples of the commercial building sites (PERMANOVA, all p < 0.05), with average dissimilarities, depending on the sampling month, ranging between 33 and 44% (mean d_BC_ range = 0.327 – 0.437; Table S7). Similar findings were associated with the residential household sites; however, three months (i.e., August, October, and November) displayed insignificant differences in community composition (PERMANOVA, all p > 0.05; Table S7). For each commercial building and residential household tap, we also estimated the pairwise dBC between flushing timepoints for all sampling months per location and used the slope of the regression fit to estimate the extent of change in community structure over flushing duration (Table S8, Figure S2 and S3). Results from time-decay relationships confirm that while flushing impacted the microbial composition of both commercial buildings and residential households, the impact was more substantial in commercial buildings. Specifically, flush duration more strongly correlated with microbial community dissimilarity at the commercial building sites and were associated with steeper slopes that were significant; however, at the residential household sites, these time-decay relationships were largely insignificant (Figure 4; Table S8). Interestingly, the linear regression fit slope of the time-decay relationship at the commercial building sites sequentially decreased in successive months, while demonstrating no significant trend at the residential household sites; this suggests that with successive month relaxation of building closures, flushing had a reduced impact on the microbial community. This is likely due to both reduced periods of stagnation as well as steady decreases in the impact of initial extended stagnation with phased re-opening, over the course of the sampling timeframe.

**Figure 4:**
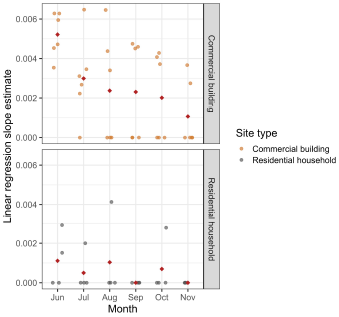
Linear regression slope estimates as determined from time-decay relationships between Bray-Curtis dissimilarity distances of any two time point combinations and flush duration between them. The analysis was performed monthly for each commercial building (orange) and residential household (grey) site, and a slope of 0 was denoted to insignificant Pearson’s correlations (p < 0.05) (see Figures S2, S3, and Table S8). The red diamond represents the mean calculated across the linear regression slope estimates.

### Microbial communities in commercial buildings became more similar to residential buildings after the relaxation of commercial building closures

Post-first flush samples (TP_2-7_) were used to assess spatial and temporal associations between microbial communities of the commercial building and residential household sites. NMDS analyses using Bray-Curtis dissimilarity distances indicated clustering of post-first flush samples by site type within each month (Figure 5A) with greater support and separation between site types observed in June, July, August, and September (PERMANOVA, R^2^ = 0.100 – 0.190, all p < 0.05; Table S9), while separation in October and November were significant but had weaker support (PERMANOVA, R^2^ = 0.021 and 0.061 for October and November, all p < 0.05; Table S9). Irrespective of the month, the microbial community composition of the commercial building and residential household sites were distinct (PERMANOVA, all p < 0.05), with average dissimilarities ranging between 35 and 46% (mean d_BC_ range = 0.350 – 0.426; Table S9). Furthermore, the microbial community composition of the commercial building and residential households sites were less dissimilar in October (d_BC_ = 0.384 ± 0.026) and November (d_BC_ = 0.384 ± 0.012) and presented lower variation with near equal dissimilarities in community composition, compared to the preceding months (Figure 5B, Table S9).

**Figure 5:**
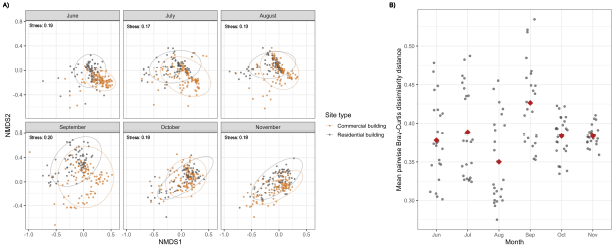
**A)** NMDS plot illustrating monthly differences in microbial community composition between TP_2-7_ samples of commercial buildings (orange) and residential households (grey) sites, using Bray-Curtis dissimilarity distances on flow cytometric data (i.e., FL-1, FL-3, FCS, and SSC). Ellipse are drawn at 95% confident limit. **B)** Mean pairwise Bray-Curtis dissimilarity distances between monthly TP_2-7_ samples of commercial building and residential household sites. The red diamond represents the averaged that were calculated across monthly mean pairwise Bray-Curtis dissimilarity distances.

To assess the impact of flow cytometry parameters in Figure 5A, we iteratively performed Bray-Curtis dissimilarity estimation and NMDS clustering using a range of combination of all four parameters (FL1, FL3, SSC, and FSC) to recapitulate the site-specific clustering followed by temporal convergence. As shown in Figure 6A, comparable observations were made when Bray-Curtis dissimilarity distances on FL1 and FL3 flow cytometric parameters were considered. Again, samples cluster by site type and presented greater separation the first four months with higher support (PERMANOVA, R^2^ = 0.145 – 0.278, all p < 0.05), when compared to October (PERMANOVA, R^2^ = 0.028, p < 0.05) and November (PERMANOVA, R^2^ = 0.095, p < 0.05; Table S10). This suggests that the observed compositional variation between the commercial building and residential household sites relate to differences in nucleic acid content distributions of flow cytometric events. Overall, the commercial building sites were associated with significantly greater HNA cell concentrations, when compared to the residential household sites (Figure 6B) (independent t-test, all p < 0.05; Table S11). These differences were particularly noticeable in June and July, where the commercial building sites were associated with HNA concentrations that were 20-fold and 10-fold greater than the residential household sites, respectively (Table S11). Bray-Curtis based dbRDA indicated that variability in microbial community composition was significantly explained by month and nine water chemistry parameters, including total chlorine, temperature, pH, conductivity, ammonium, nitrite, Fe^54^, Mn^55^, and Zn^66^ (permutation test for Bray-Curtis dbRDA, all *p* < 0.01; Table S12A). The dbRDA plot presented in Figure 6C, showed clustering of samples by month that explained 27% of the variation in microbial community composition based on variance partitioning analysis (adj. R^2^ = 0.273; Table S12B). Among the water chemistry parameters, total chlorine, ammonium, and conductivity were identify as primary water chemistry parameters driving change in microbial community composition between months (variance partitioning analysis, adj. R^2^ = 0.093, 0.098, and 0. 069; Table S12B). Of these, total chlorine and ammonium explained most of the variation and were present at lower concentration in June and July (Table S1). Furthermore, these water chemistry parameters had significant correlations with HNA concentrations. Specifically, total chlorine strongly negatively correlated with HNA concentrations (*r*(34) = - 0.77, *p* < 0.05), while ammonium moderately negatively correlated with HNA concentrations (*r*(34) = - 0.40, *p* < 0.05). These findings suggesting that temporal changes in residual chlorine and ammonia concentration, both impacted by stagnation in a chloraminated system, likely drive change in nucleic acid content distributions of the microbial communities in commercial building and residential household sites.

**Figure 6:**
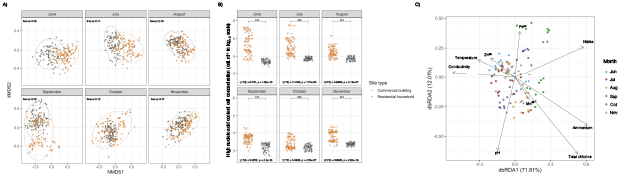
**A)** NMDS plot illustrating differences in microbial community composition between TP2-7 samples of commercial buildings (orange) and residential households (grey) sites, using Bray-Curtis dissimilarity distances on flow cytometric data (i.e., FL-1 and FL-3). Ellipse are drawn at 95% confident limit. **B)** High nucleic acid content cell concentration in cells per ml (determined in triplicate) for TP2-7 samples that were collected monthly in the commercial building (orange) and residential household (grey) sites. Differences between the commercial building and residential household sites were assessed using independent t-test and significance are indicated with a significant bar, with 0.01 ≤ p < 0.05 (*), 0.001 ≤ p < 0.01 (**), p < 0.001 (***), and not significant (ns). **C)** Bray-Curtis distance-based redundancy analysis (dbRDA) biplot illustrating the relationship between sampling month, water chemistry parameters and microbial community composition of TP7 samples of the commercial building and residential household sites, respectively. Water quality parameters that significantly explained variation in community composition based on permutational ANOVA for Bray-Curtis dbRDA (p < 0.01) are shown as black arrows and the percent variation explained by each axis is shown in parenthesis.

Thus, our study demonstrates that COVID-19 related building closures in March 2020 not only impacted water demand but also microbial community composition in commercial building sites as compared to residential household sites. While this impact on the drinking water microbial community of commercial building sites could not be mitigated through short-term flushing regimes (30 minutes) following phased reopening, the microbial community demonstrated gradual recovery over a period of months as indicated by their convergence towards the microbial communities at the residential household sites. This suggests that sustained and gradual increase in water demand may play a more important role in the recovery of building plumbing-associated microbial communities as compared to short-term flushing after extended periods of altered water demand that result in reduced flow volumes.

## Data Availability

The raw FCS files have been deposited in FlowRepository and are publicly available under accession number FR-FCM-Z4SR.

