## Supplementary figures and images for "Systematic recovery of building plumbing-associated microbial communities after extended periods of altered water demand during the COVID-19 pandemic"

### Supplemental Figure S1

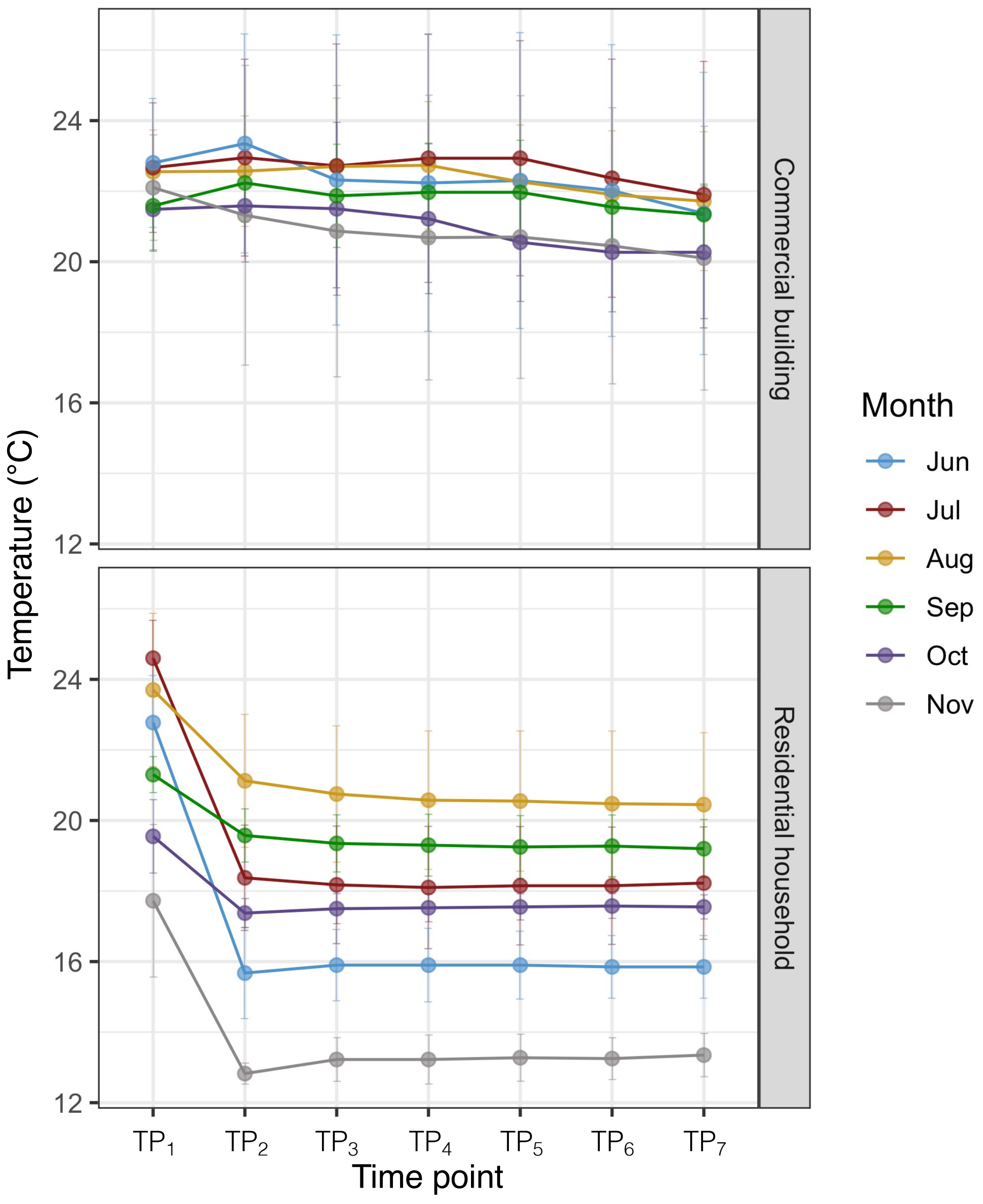

### Supplemental Figure S2

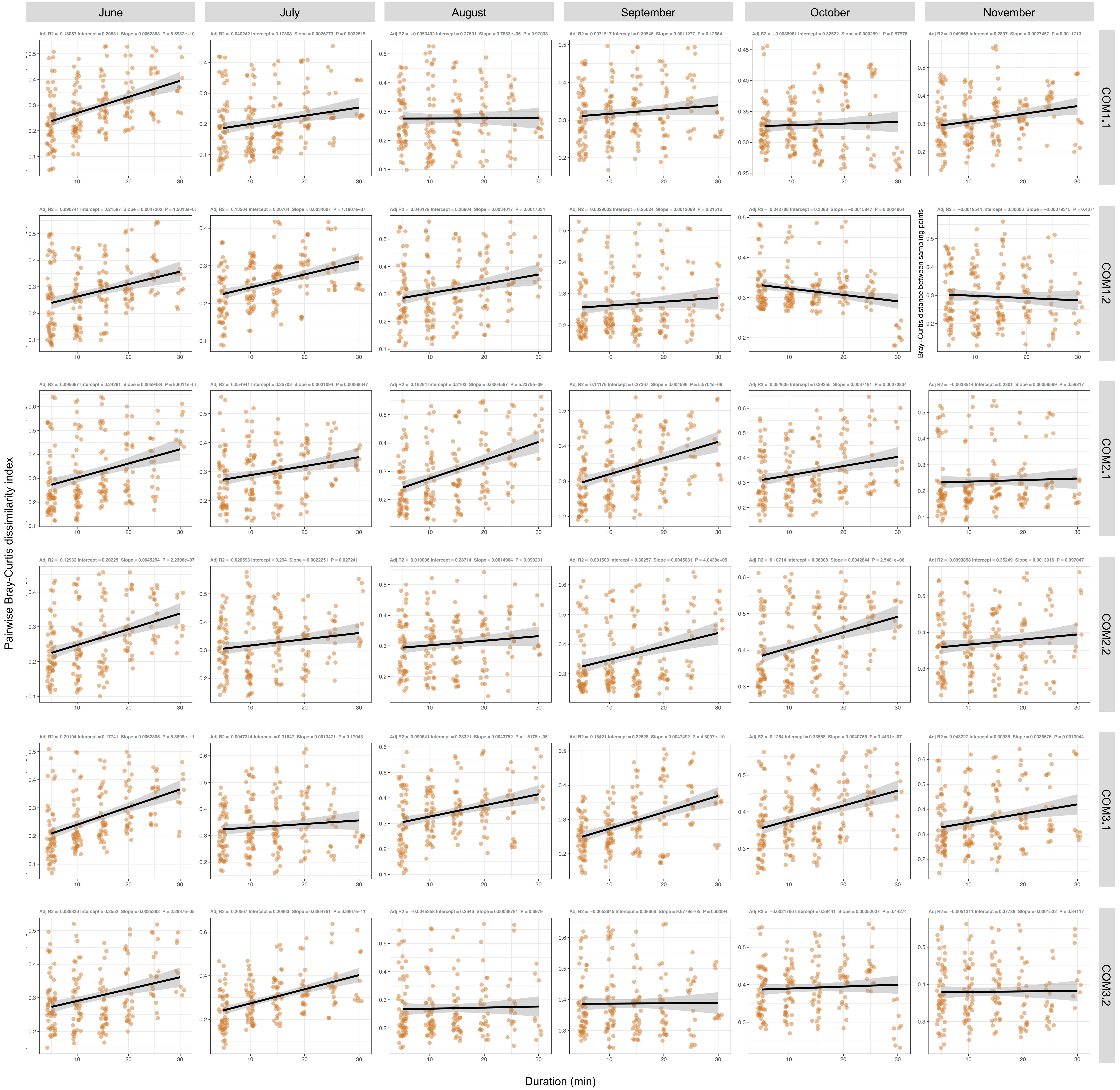

### Supplemental Figure S3

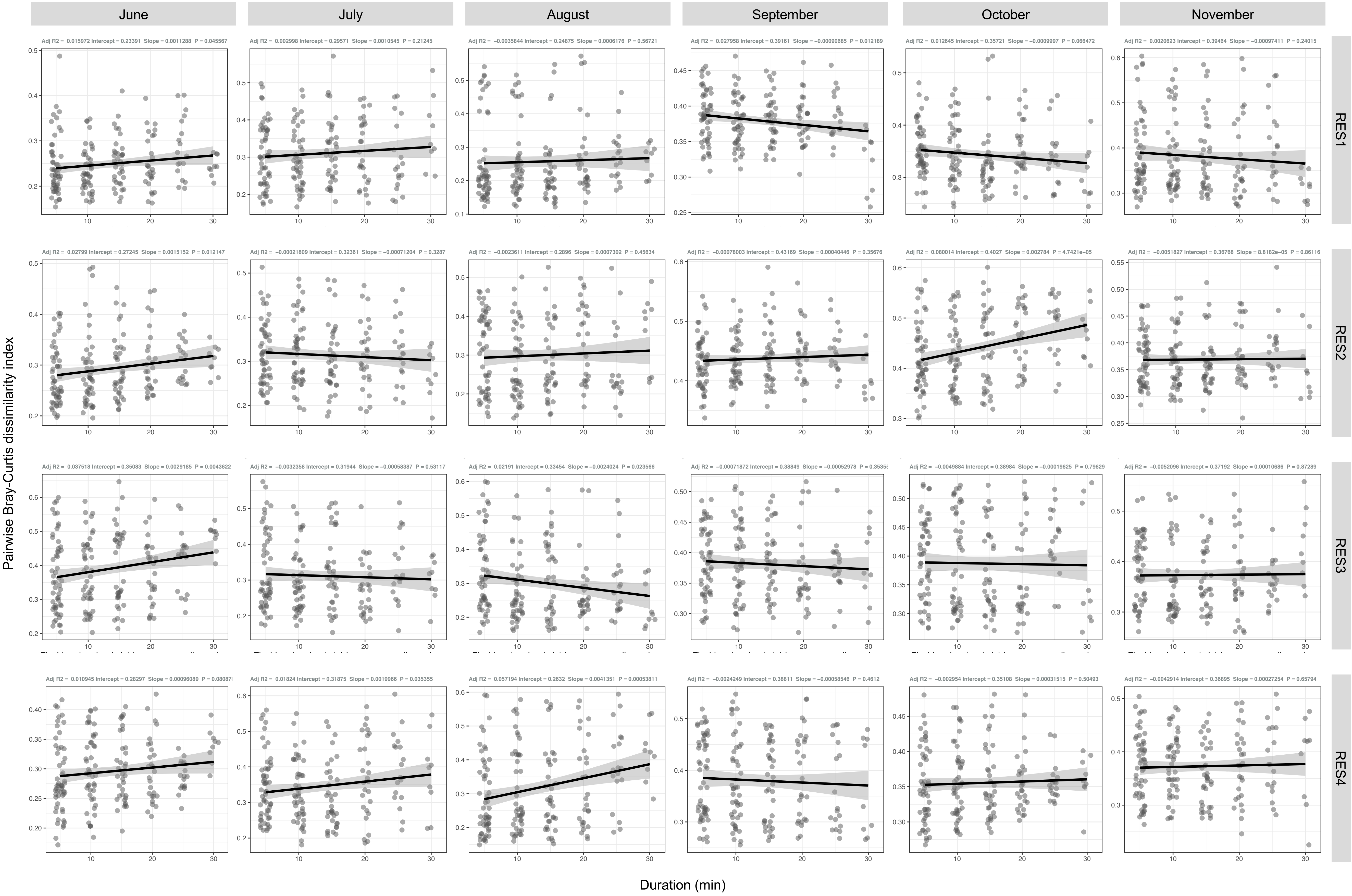
